# Impact of Remote Monitoring on Clinical Outcomes in Defibrillator Patients During the COVID-19 Pandemic: An Interrupted Time Series Analysis

**DOI:** 10.1101/2024.10.22.24315956

**Authors:** Sandeep Chand, Barbara Torres, David Vickers, Tara A Whitten, Andrew Mardell, Glen Sumner

**Author notes:** ***Corresponding author:*** Sandeep Chand, Community Health Sciences, Cumming School of Medicine, University of Calgary, Alberta, Canada.

## Abstract

**Background:** Remote monitoring (RM) has become the standard of care in many Cardiac Implantable Electronic Device (CIED) clinics across North America and Europe. Some clinics are even adopting an alert-based, RM-only strategy for select patients. However, it remains unclear whether RM, compared to usual non-remote care, impacts the total number of CIED clinic visits, emergency department visits, hospitalizations, and all-cause mortality. Because such outcomes and their relationship to RM may be magnified in the presence of a global COVID-19 pandemic, we aimed to perform an interrupted time series analysis to observe trends in these outcomes in response to the declaration of the COVID-19 pandemic in patients with implantable cardioverter-defibrillators (ICD).

**Methods:** In this retrospective study, we utilized existing electronic provincial databases maintained by Alberta Health Services (AHS) and Alberta Health (AH) to determine CIED visits, emergency room visits, cardiovascular (CV) hospitalizations, and all-cause mortality. We performed Interrupted Time Series (ITS) analysis to compare outcome trends in ICD-patients with and without RM in Alberta, both during the COVID-19 pandemic and the pre-pandemic period. We defined the time-period of the pandemic as March 17, 2020, to July 17, 2021. Pre-pandemic was defined as March 17, 2018, to March 16, 2020. We compared best model fits using the Akaike Information Criterion (AIC), selecting the model with the lowest AIC for each outcome. The best-fitting models were plotted. Outcomes between RM and non-RM groups were compared using regression models, with differences reported using 95% confidence intervals.

**Results:** The CIED population consisted of 6,183 ICD patients from March 17, 2018, to July 17, 2021. Of these, 2,989 (48.3%) had access to RM. Our study found that access to virtual consultations sharply increased at the onset of the pandemic in both cohorts, though this trend was significantly higher in the RM group. Conversely, a sharp decline in in-person visits was observed for RM patients. Compared to those without RM, patients with RM showed no significant differences in all-cause mortality, hospitalizations, or emergency room visits, and these trends were not impacted by the COVID-19 pandemic.

**Conclusion:** In ICD patients with and without RM, the number of virtual consultations increased while in-person visits decreased during the pandemic. However, no significant changes in the trends of cardiovascular hospitalizations, emergency room visits, or all-cause mortality were observed in either group during this period. This suggests that RM did not significantly impact key health outcomes for ICD-patients during the pandemic in Alberta.

## 1.0 Background

SARS-CoV-2 infection was declared a world pandemic by the World Health Organization (WHO) on 11 March 2020.^1^ The COVID-19 pandemic triggered a significant shift in healthcare delivery, prompting policymakers and healthcare providers to rapidly transform how patients are monitored and treated. Among the various adaptations in healthcare, remote monitoring (RM) has emerged as a pivotal strategy, particularly for patients with cardiovascular implantable electronic devices (CIEDs).^2^

RM allows timely transmission of CIED-stored diagnostic data to a central server and analysis of this data by a trained nurse clinician. There is growing evidence that communication of the RM data to a heart rhythm specialist physician may improve quality of care.^3^ Further, prompt detection of arrhythmia and of heart failure deterioration may decrease emergency department visits, hospital admissions and hospital length of stay.^4-5^ In practice, there are now selective CIED clinics that have adopted an “alert-based” or RM-only care with CIED clinic visits reserved only for use on an urgent, ad hoc basis.^.6^ Multiple guidelines have recommended the adoption of a RM strategy for patients with CIEDs across the globe.^7-8^ Though there is evidence that RM helps in reducing mortality and healthcare resource utilization, it is unclear whether it is still impactful and effective during the COVID pandemic. Thus, we intended to use administrative data from device clinics in Alberta, Canada to conduct an interrupted time series (ITS) analysis to determine the effect of the COVID-19 pandemic on health care utilization and cardiovascular (CV) outcomes in patients with CIEDs. We focused our analysis on the population of people with defibrillators both with and without RM.

## 2.0 Methods

In this retrospective cohort study with an ITS analysis, we used aggregated and anonymized administrative outcome data maintained by Alberta Health Services (AHS) and Alberta Health (AH). The data provided comprehensive, cumulative and longitudinal data for all known CIED patients in Alberta, Canada. This jurisdiction is a publicly financed, universal access health system with almost all provincial residents enrolled in the provincial health plan. Outcomes were ascertained through linkage of CIED patients’ de-identified personal health number to the outcome databases using the International Classification of Diseases, Tenth Revision (ICD-10).^9^ We defined the pre-intervention study period as March 17, 2018 to March 16, 2020 with the intervention to be a specific 16 months’ time period of the COVID-19 pandemic in Alberta: March 17, 2020 to July 17, 2021. We chose an interrupted time series analysis as a sensitive means to detect any potential effect in trends of our outcomes of interest: CIED visits, emergency room visits, CV hospitalizations, and all-cause mortality. This research study was approved by University of Calgary’s Conjoint Health Research Ethics Board with waiver of informed consent.

### 2.1 Study population

We analyzed all CIED clinics patients 18 years older in Alberta with defibrillators who accessed CIED clinics between March 17, 2018, and July 17, 2021 using the CIED province-wide clinic database. We divided ICD-patients into RM and non-RM (Fig 1). Patient’s age, biological sex, clinic location, RM status, emergency department visits, comorbidities, CIED visits (in-person visits, or virtual consultations) and month of death were considered for the study. PaceArt database was used to identify the patients with RM status. Comorbidities were identified using the ICD-10 codes.

**Fig. 1:**
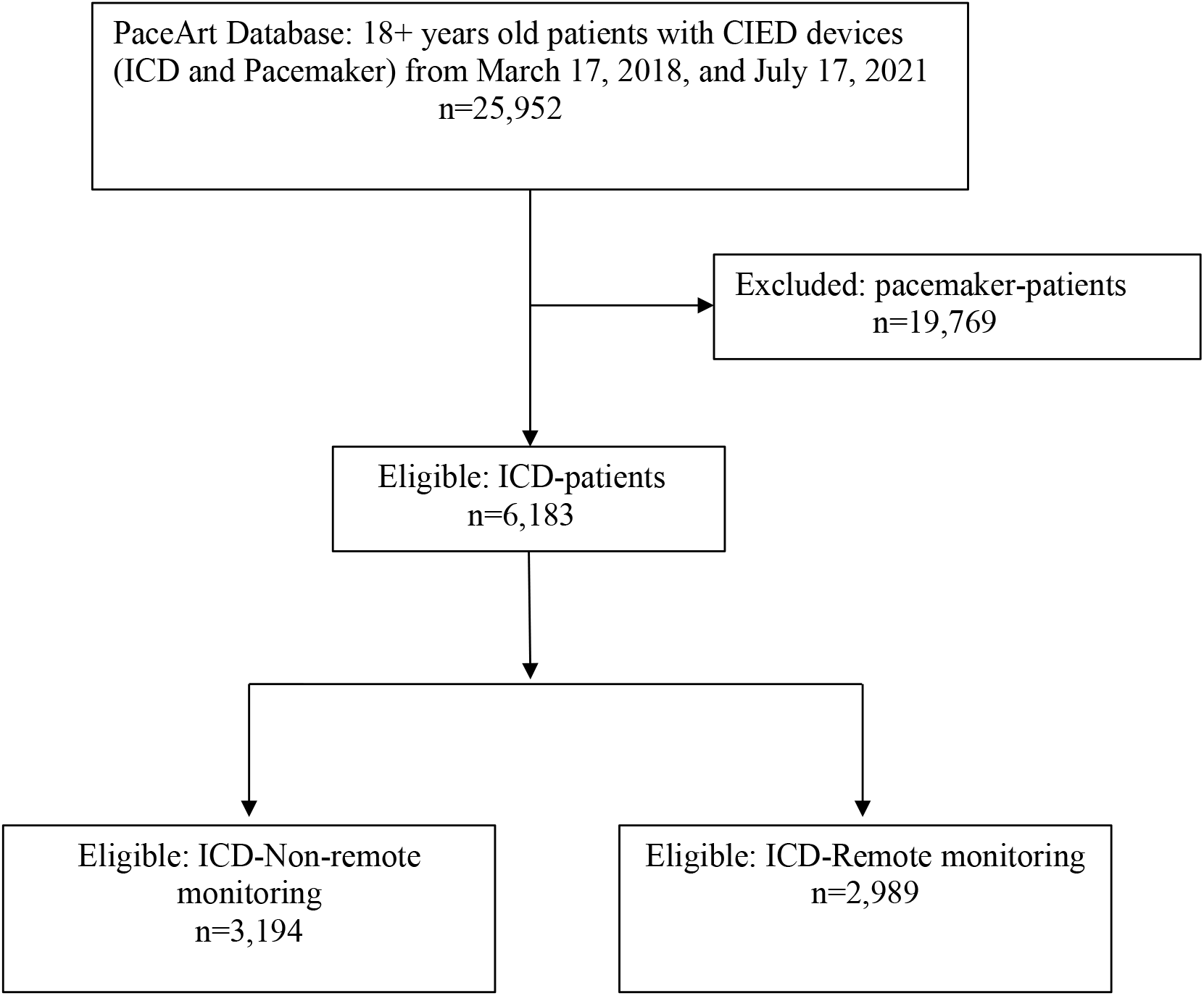
Inclusion/exclusion flow diagram This diagram illustrates the inclusion and exclusion criteria used to create cohorts. CIED= Cardiac implantable electronic device; ICD= Implantable cardioverter-defibrillator

### 2.2 Outcomes

We compared trends in overall visits (in-person visits and virtual consultations), ED visits, CV-hospitalizations, and all-cause mortality in patients with defibrillators with/without RM identified using anonymized patient identification numbers. The natural exposure was the initial 16 months period of the pandemic as defined above. Thus, after 3 years, outcome data was determined with secular trend pre-pandemic, and then analyzed during the pandemic for ICD-patients with/without RM. More details on measurement of the outcomes are provided in the supplement **(Supplement)**.

#### 2.2.1 Overall visits

We assessed the volume of overall visits by combining in-person visits and virtual consultations from physician claims dataset from March 17, 2018, and July 17, 2021. We defined virtual visits as any interaction between patients and CIED clinic members (nurse clinicians, physicians) that was not conducted in person.

#### 2.2.2 Emergency department visits

ED visits were the visits that resulted in discharge from the emergency department, due to heart failure, arrhythmias, stroke, or MI that involved physician care in an emergency room or urgent care setting. CV related ER visits were identified using the national ambulatory care reporting system (NACRS) database from March 17, 2018, and July 17, 2021.

#### 2.2.3 CV Hospitalizations

CV hospitalizations included admissions for heart failure (HF), arrhythmias, stroke, or myocardial infarction (MI). We identified hospitalization data using the discharge abstract databas**e (**DAD) from March 17, 2018, and July 17, 2021.

#### 2.2.4 All-cause mortality

All-cause mortality was defined as the deaths of active CIED clinic patients. Mortality data was identified using the vital statistics database from March 17, 2018, and July 17, 2021.

### 2.3 Statistical Analyses

The dichotomous outcomes were measured as a proportion of the total ICD-patients with RM vs. non-RM in monthly intervals between March 17, 2018 to July 17, 2021. Each outcome was plotted on a separate graph based on the best fit of data. Outcome comparisons between RM vs. non-RM were described as differences with 95% confidence intervals using appropriate statistical techniques for discrete variables.

To describe the trends in our outcome variables, over time, amongst RM and non-RM patients we employed three different regression models:

1. a standard linear regression model with monthly time points as the sole independent variable, 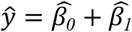 *time*;
2. an interrupted time series (ITS) model with March 2020 included as a “dummy” variable to represent healthcare responses to COVD-19, 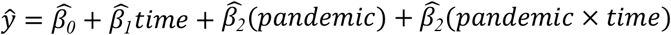; and
3. a segmented regression model, where an outcome is modelled as two separate linear functions on either side of a breakpoint (or “hinge”, *h*). Here, h was automatically chosen by using the R package *segmented* (version 1.6-4) to produce the fitted model, 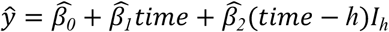, where *I*_*h*_ = *0* when *time* ≤ *h* and *I*_*h*_ = *1* when *time* > *h*.

For reach model, *β*_*0*_ represents the baseline level at time = 0, *β*_*1*_ is the slope of an outcome per month (representing the underlying trend before March 2020); in the ITS model, *β*_*2*_ is a level change following March 2020, and *β*_*3*_ indicates the slope change following March 2020; *β*_*2*_ in the segmented model indicates the slope following a hinge point, h.

We included ITS and segmented regression models because changes in intercept and/or slope from March 2018 to July 2021 were used to test causal hypotheses about the influence of the responses to COVID-19 on the outcomes listed above. To find out which of the three models best described the trends in PaceArt participants, we compared them using their Akaike information criterion (AIC). The model with the lowest AIC was deemed the best-fit to the available data. All regression analyses used R statistical software (version 4.2.3).

## 3.0 Results

### 3.1 Cohort Characteristics

We identified a total of 6183 adults with a defibrillator between March 17, 2018, and July 17, 2021.We observed that 48% were using RM. The proportion of patients with cardiac resynchronization therapy (CRT) was 29% in non-RM and 28% in RM groups. Median age was 65 years (non-RM) and 67 years (RM), with approximately 22% females in both groups, predominantly from urban CIED clinics (table 1). We observed 19% of non-RM patients and 11% of RM patients had Coronary and peripheral vascular disease (CPVD) (table 1). The cohorts were well matched with respect to the baseline of pre-existing conditions. However, HF prevalence at baseline was higher in the non-remote group (27% vs 18%) (table 1).

**Table 1:** Baseline population characteristics for patients with ICD.

| Characteristic Monitoring Type | Non-Remote Monitoring | Remote Monitoring |
| --- | --- | --- |
| **Volume n (%)** | 3,194 (52%) | 2,989 (48%) |
| **CRT, n (%)** |  |  |
| CRT | 934 (29%) | 824 (28%) |
| Non-CRT | 2,260 (71%) | 2,165 (72%) |
| **Age, Median (Range)** | 65 (18-93) | 67 (18-94) |
| **Female n (%)** | 697 (22%) | 667 (22%) |
| **Location of clinics n (%)** |  |  |
| Urban clinics | 3,093 (97%) | 2,846 (95%) |
| Rural clinics | 91 (3%) | 143 (5%) |
| **Comorbidities n (%)** |  |  |
| Coronary and peripheral vascular disease (CPVD) | 608 (19%) | 321 (11%) |
| Ischemic cardiomyopathy (ICM) | 991 (31%) | 761 (25%) |
| Heart Failure | 868 (27%) | 544 (18%) |
| Stroke | 59 (2%) | 42 (1%) |
| Atrial fibrillation | 503 (16%) | 374 (13%) |
| Supraventricular tachycardia (SvT) | 63 (2%) | 53 (2%) |
| Ventricular arrhythmia (VA) | 493 (15%) | 343 (11%) |
| Cardiac arrest | 265 (8%) | 91 (3%) |
| Hypertension | 861 (27%) | 709 (24%) |
| Diabetes | 744 (23%) | 572 (19%) |
| Dyslipidemia | 162 (5%) | 149 (5%) |
| Renal disease | 124 (4%) | 112 (4%) |

CRT= Cardiac resynchronization therapy; CPVD= Coronary and peripheral vascular disease; Ischemic cardiomyopathy; ICM= Ischemic cardiomyopathy; SvT=Supraventricular tachycardia; VA= Ventricular arrhythmia

### 3.2 Overall Visits trends (in-person visits and virtual)

Patients with non-RM experienced a declining trend in overall visits beginning in March 2018, decreasing from approximately 306 per 1,000 patients to 85 visits per 1,000 patients (Fig.2) (slope = -9.014, 95% CI: -10.366, -7.662) (supplement: eTable 1). In July 2021, the number of visits had dropped to approximately 74 per 1,000 patients (Fig.2), with a slower decline observed during this period (slope = -1.234, 95% CI: -2.586, 0.119) (supplement: eTable 1).

**Fig 2:**
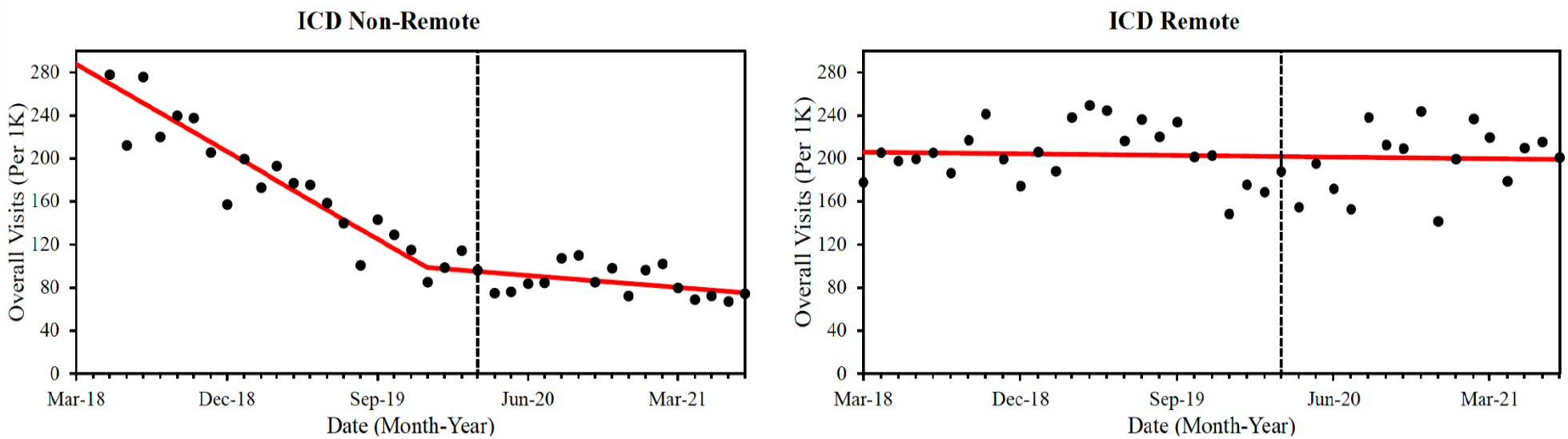
Best-fitting models (red line) for the total number of overall visits (per 1,000 patients) in both the Non-Remote and Remote groups (black dots) by month and year. The vertical dashed line indicates the beginning of the pandemic period, March 17, 2020. ICD= Implantable cardioverter-defibrillator.

In contrast, no interruption in the trend was observed for patients with RM (supplement: eTable1). These patients showed a slight downward trend, with visits decreasing from approximately 205 per 1,000 patients in March 2018 to approximately 201 per 1,000 patients in July 2021 (Fig.2).

#### 3.2.1 In-person visits

In non-RM patients, in-person visits declined from approximately 248 per 1,000 patients to around 35 per 1,000 patients in March 2020 (Fig.3) (slope = -6.187, 95% CI: -7.833, -5.801) (supplement: eTable 1). Following March 2020, the trend remained nearly stable (slope = 0.364, 95% CI: -1.502, 2.230) (supplement: eTable 1), with in-person visits at approximately 32 per 1,000 patients (Fig. 3).

**Fig 3:**
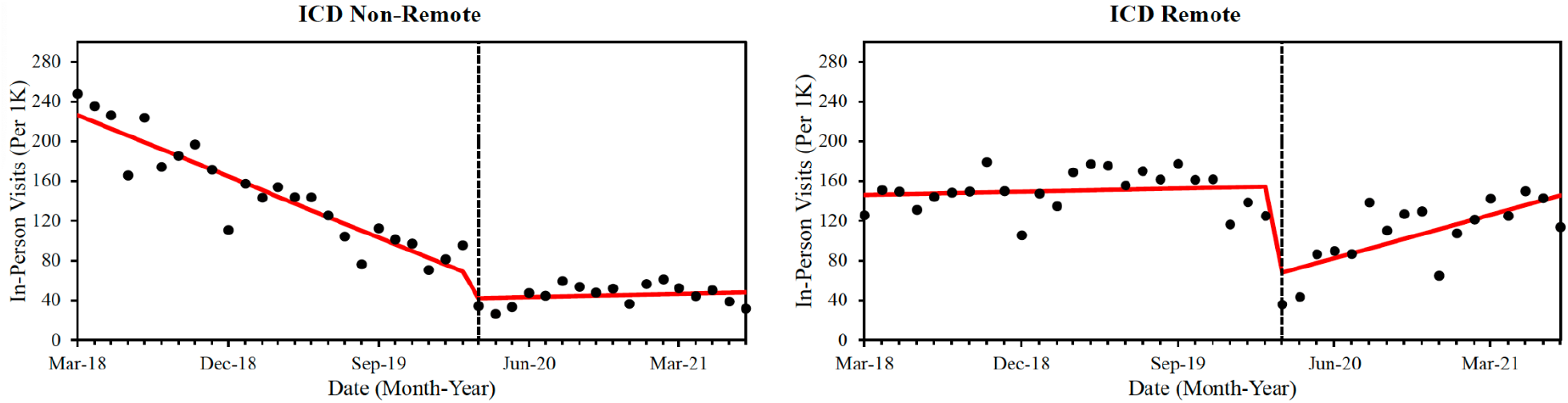
Best-fitting models (red line) for the total number of in-person visits (per 1,000 patients) in both the Non-Remote and Remote groups (black dots) by month and year. The vertical dashed line indicates the beginning of the pandemic period, March 17, 2020. ICD = Implantable cardioverter-defibrillator.

In contrast, RM patients showed a mild increase in visit trends before the pandemic, but this significantly decreased to 36 per 1,000 patients in March 2020 (Fig. 3) (slope = 0.235, 95% CI: - 1.155 to 1.625) (supplement: eTable 1). This was followed by a rebound to 114 per 1,000 patients in July 2021 (Fig. 3) (slope = 4.947, 95% CI: 2.231 to 7.658) (supplement: eTable 1).

#### 3.2.2 Virtual consultations

In non-RM patients, the trend for virtual consultations decreased from approximately 58 per 1,000 patients to 19 per 1,000 patients before pandemic (Fig. 4) (slope = -1.860, 95% CI: -2.265, -1.456) (supplement: eTable 1). In March 2020, consultations briefly increased to 62 per 1,000 patients but resumed a slower decline with 43 visits per 1,000 patients in July 2021 (Fig. 4) (slope = -1.537, 95% CI: -2.280 to -0.793) (supplement: eTable 1).

**Fig 4:**
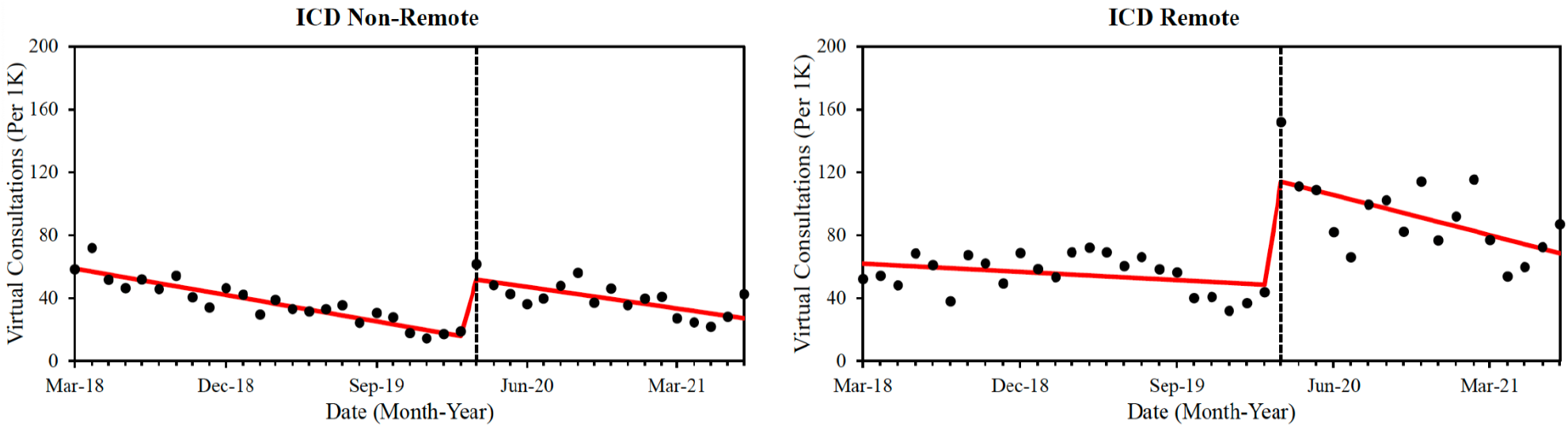
Best-fitting models (red line) for the total number of virtual consultations (per 1,000 patients) in both the Non-Remote and Remote groups (black dots) by month and year. The vertical dashed line indicates the beginning of the pandemic period, March 17, 2020. ICD = Implantable cardioverter-defibrillator.

RM patients experienced a downward trend in virtual consultations pre-pandemic, decreasing from 52 per 1,000 patients in March 2018 to 22 per 1,000 patients in February 2020 (Fig.4) (slope = -0.582, 95% CI: -1.525 to 0.362) (supplement: eTable 1). Consultations then surged significantly to 152 per 1,000 in March 2020 before decreasing again to 87 per 1,000 patients in July 2021 (Fig. 4) (slope = -2.843, 95% CI: -4.687 to -0.998) (supplement: eTable 1).

### 3.3 Emergency department visits

In the non-RM group, ED visits showed a downward trend before January 2021, decreasing from approximately 2 per 1,000 patients in March 2018 to 0.3 per 1,000 patients in January 2021 (Fig. 5) (slope = -0.083, 95% CI: -0.101, -0.065) (Supplement: eTable 2). After January 2021, ED visits increased to 1 per 1,000 patients in July 2021 (Fig. 5) (slope = 0.119, 95% CI: -0.091, 0.328) (Supplement: eTable 2).

**Fig 5:**
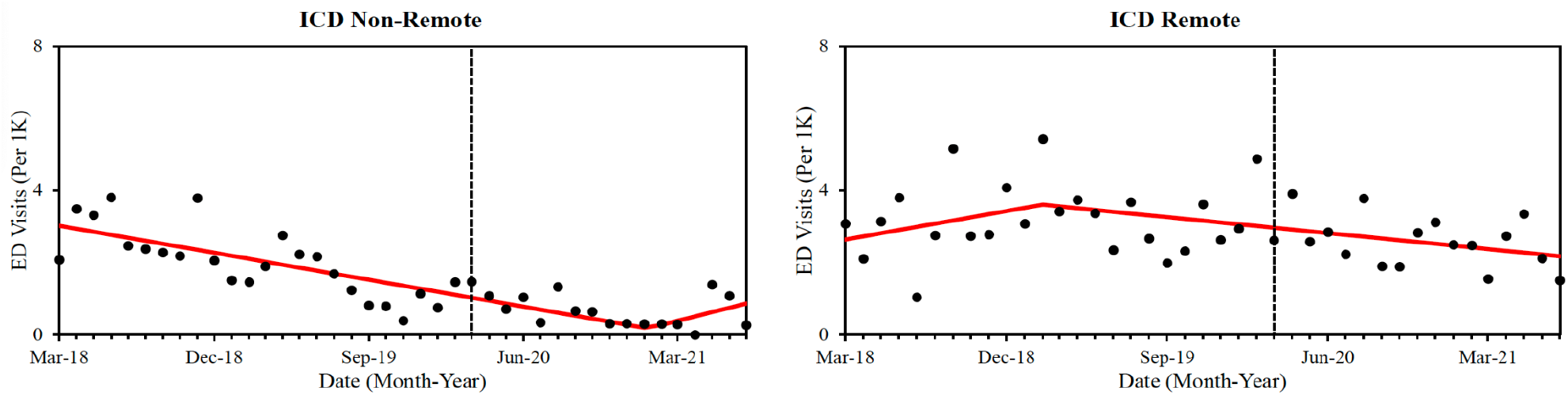
Best-fitting models (red line) for the total number of ED visits (per 1,000 patients) in both the Non-Remote and Remote groups (black dots) by month and year. The vertical dashed line indicates the beginning of the pandemic period, March 17, 2020. ICD = Implantable cardioverter-defibrillator.

In the RM group, ED visits increased from 3 ED visits per 1,000 patients in March 2018 to 5 ED visits per 1,000 patients in February 2019 (Fig.5) (slope = 0.088, 95% CI: -0.082, 0.258), and then decreased to 1.5 per 1,000 patients in July 2021 (Fig.5) (slope = -0.049, 95% CI: -0.087, 0.012) (Supplement: eTable 2).

### 3.4 CV hospitalizations

In the non-RM group, CV hospitalizations slightly decreased pre-pandemic, from 4 to 2 per 1,000 patients (Fig. 6) (slope = -0.191, 95% CI: -0.274 to -0.109) (Supplement: eTable 3), then stabilized post-pandemic at around 1 per 1,000 patients in July 2021 (Fig. 6) (slope = 0.001, 95% CI: -0.029 to 0.333) (Supplement: eTable 3).

**Fig 6:**
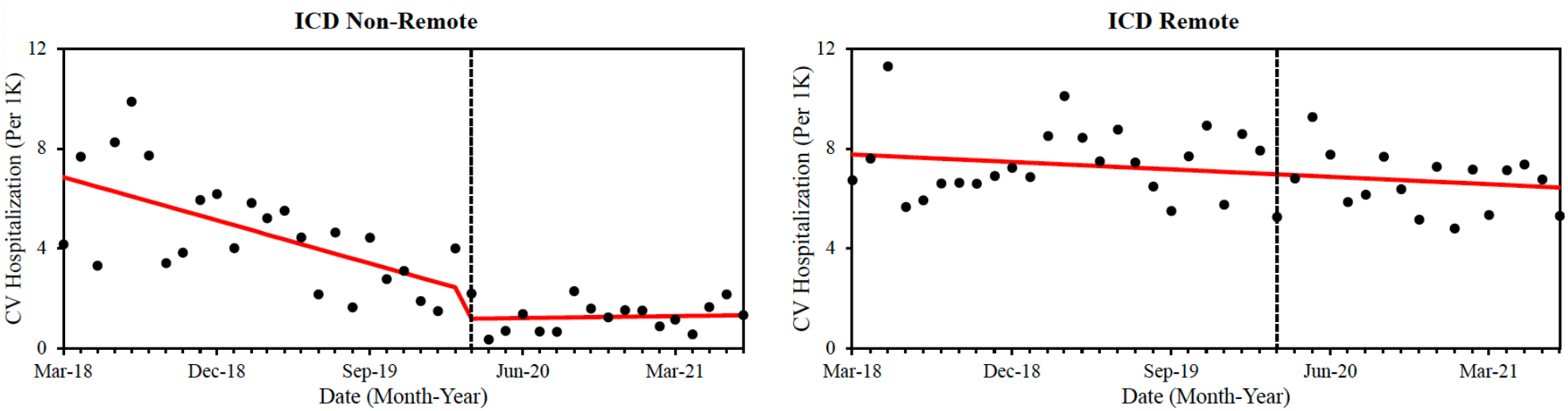
Best-fitting models (red line) for the total number of CV hospitalizations (per 1,000 patients) in both the Non-Remote and Remote groups (black dots) by month and year. The vertical dashed line indicates the beginning of the pandemic period, March 17, 2020. ICD = Implantable cardioverter-defibrillator.

The CV hospitalizations trend in the RM group was not interrupted during the study period. It decreased from 7 per 1,000 patients in March 2018 to 5 per 1,000 patients in July 2021 (Fig. 6)

### 3.5 All-cause mortality

We found no interruption in the trend of all-cause mortality rates per 1,000 patients in either group during COVID-19 pandemic (Fig. 7 and Supplement: eTable 4). In non-RM patients, all-cause mortality demonstrated a significant downward trend since March 2018, decreasing from approximately 5 per 1,000 patients to 2 per 1,000 patients in July 2021 (Fig. 7).

**Fig 7:**
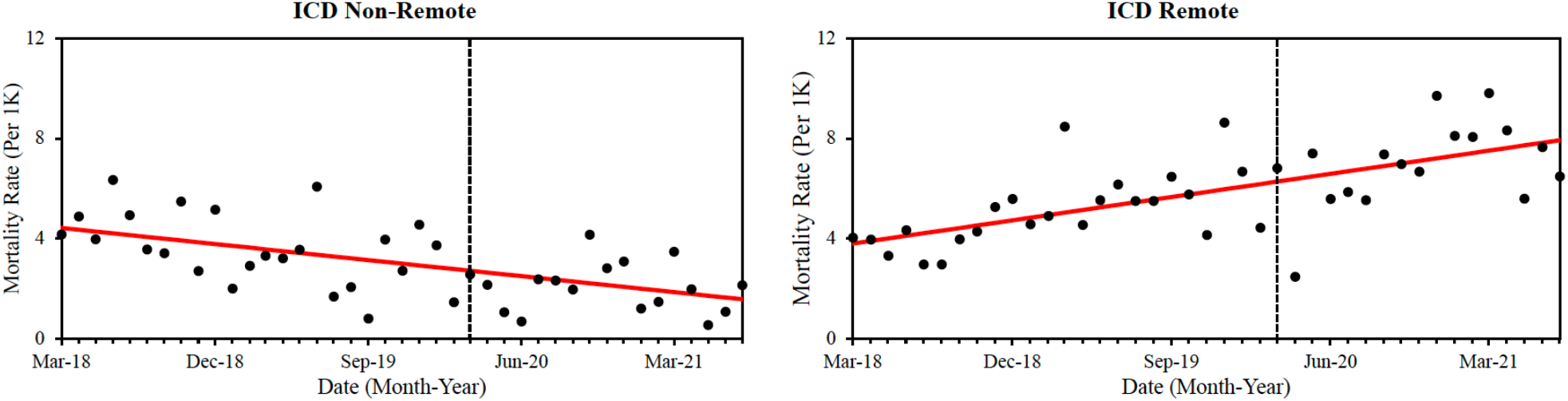
Best-fitting models (red line) for the total number of all-cause mortality (per 1,000 patients) in both the Non-Remote and Remote groups (black dots) by month and year. The vertical dashed line indicates the beginning of the pandemic period, March 17, 2020. ICD= Implantable cardioverter-defibrillator; 1K= 1,000; Mon= Month; Yr= Year.

In contrast, the RM group exhibited a significant upward trend in all-cause mortality over the same period, increasing from approximately 4 per 1,000 patients in March 2018 to 8 per 1,000 patients in July 2021 (Fig. 7).

## 4.0 Discussion

In our study, RM patients had higher overall visits than non-RM patients through the study-period (Fig.2). The COVID-19 pandemic did not significantly impact overall visits and mortality trends in either group. Notably, there was a rise in virtual consultations for both cohorts coinciding with the pandemic’s declaration, particularly in the RM group. This increase in virtual care consultations for RM patients may be attributed to uninterrupted remote device transmissions despite limited clinic access during the pandemic’s early phase. ^10^ Moreover, both scheduled and unscheduled device transmissions necessitate virtual consultations from CIED clinic staff.^10^ Conversely, non-RM patients needed to initiate contact for virtual consultations in the absence of scheduled clinic appointments.

Our study found no significant benefit of remote monitoring (RM) in reducing ED visits and virtual consultations, consistent with findings from a recent systematic review and meta-analysis study.^11^ Moreover, RM did not demonstrate a substantial reduction in cardiovascular hospitalizations compared to non-RM patients, which contrasts with the findings of the systematic review and meta-analysis.^11^

Thus, the COVID-19 pandemic did not significantly impact the trends in overall visits, ED visits, hospitalizations, and mortality in the RM group and the reason remains unclear. However, RM may have facilitated virtual consultations from device nurse clinicians despite limited in-person access. This hypothesis is supported by the noted increase in virtual consultations for ICD patients with RM (Fig 4). In the non-RM group, ED visit rates increased modestly one-year post-pandemic. This may be due to lower virtual care and limited in-person access.

Overall, no significant pandemic-related changes in clinical outcomes were observed especially on RM group. Alternatively, patient access to care from clinicians was maintained through increased virtual care during the pandemic. Consequently, the overall rise in virtual access for both ICD patient groups likely prevented interruption of the declining trend in the clinical outcomes.

Due to the specificity of our approach, we were unable to find directly comparable studies in the existing literature. However, it is worth noting the study conducted by Nagy B. et al. (2021), observed a reduction in hospital/in-clinic visits during the lockdown period as compared from the pre-pandemic time in RM patients with CIEDs in Hungary.^12^ Additionally, Nagy et al. noted that the trend in hospital visits rebounded following the lifting of lockdown restrictions.^12^ This finding does not completely support our result as we observed a decrease in the trend of in-person visits for a limited time period of a few months after which the trend started increasing again. However, it is important to emphasize that Nagy et al.’s study had a relatively small sample size of 85 patients in their analysis.^12^

Another study by Vincenzo R. et al. (2021) reported a significant increase in unplanned hospitalizations among patients with CIEDs in Campania during the pandemic year compared to the pre-pandemic period.^13^ This increase was independent of whether or not patients were managed through RM or non-RM care, nor was it affected by their ICD status.^13^ That study found no statistically significant difference in ED visits between the pre-pandemic and pandemic periods.^13^ These findings are not entirely consistent with ours, which may be attributed to several factors such as different patient populations and timeframe considered for the reasearch.^13^ Additionally, the study did not sufficiently account for the impact of RM versus non-RM status on hospitalizations and ED visits, particularly over the shorter timeframe analyzed.^13^

Our study has limitations. Firstly, we did not control for potential confounding factors, which could have influenced our results. The use of propensity score matching, based on baseline characteristics, could have provided more accurate and adjusted outcomes. Secondly, though we considered the CIED clinic location in baseline characteristics, we did not identify the location of patients (rural versus urban) which might have contributed to the lack of significant changes in some primary outcomes. Thirdly, we did not exclude for patients who may have switched between remote and non-RM during the study period. This lack of control over “switchers” could have introduced additional variability and impacted the results, potentially diluting the observed effects of RM on the study’s outcomes. Lastly, the misclassification of patients due to incorrect ICD-10 coding may have affected the accuracy of some of our outcome measures. Thus, due to these limitations, the generalizability of our findings is reduced.

Our study has several strengths. First, the quasi-randomized nature of the design with the non-RM as a control group in a similar population gives an element of internal validity/strength to the study. Second, we included a large sample size of 6,183 adults which allowed us to detect subtle trends contributing to the robustness of our results. Third, the extended study period, which spans several years, provided a comprehensive overview of trends over time. Fourth, our study accounted for a diverse patient population, including individuals with various comorbidities and different backgrounds, making our findings reflective of the broader population.

We believe that future investigations should focus on assessing various factors such as geographical location, comorbidities, socio-economic status, and other relevant variables to better understand why there was no observed decrease in the overall clinical outcomes within the RM group as compared to the non-RM group. Additionally, another study could address the limitations of our research by employing a matched cohort design to validate the results of our study. Such an approach would enable a more robust evaluation of the impact of RM and help to clarify the underlying reasons for the observed trends.

## 5.0 Conclusion

CIED defibrillator patients with RM demonstrated an increase in virtual consultations compared to non-RM patients during the COVID-19 pandemic. In the non-RM group, a modest trend toward increased ED visits was associated with relatively less access to virtual CIED clinic care. Overall, trends in total visits, ED visits, CV hospitalizations, and all-cause mortality were not interrupted during the COVID-19 pandemic in patients with RM. Similarly, the trend in all-cause mortality in non-RM patients was also unaffected during the COVID-19 pandemic.

## Data Availability

The datasets generated and analyzed during the current study are not publicly available due to confidentiality agreements with Alberta Health Services (AHS) and Alberta Health (AH). However, the data can be made available upon request by the journal, subject to approval and compliance with confidentiality requirements. Aggregate data and summary statistics are included within the manuscript.

